# Comprehensive genetic analysis of *STRC* variants in hereditary hearing impairment using long-read sequencing

**DOI:** 10.1101/2024.11.05.24316795

**Authors:** Cheng-Yu Tsai, Yue-Sheng Lu, Yu-Ting Chiang, Ming-Yu Lo, Pei-Hsuan Lin, Shih-Feng Tsai, Chuan-Jen Hsu, Pei-Lung Chen, Jacob Shu-Jui Hsu, Chen-Chi Wu

## Abstract

**Background:** Sensorineural hearing impairment (SNHI) is a common disorder with a significant genetic component. Genetic testing for SNHI often involves next-generation sequencing (NGS), but SNHI-related pathogenic *STRC* variants cannot be directly addressed by conventional NGS due to the complex genomic scenario derived from large genomic rearrangements and a highly homologous pseudogene. Long-read sequencing (LRS) offers an unprecedented resolution to these challenges.

**Methods:** We developed a comprehensive workflow that integrates the PacBio-based LRS approach with marker-mediated refinements to effectively address pseudogene contamination. This methodology was applied to analyze the *STRC* gene in a cohort of 100 unrelated Taiwanese patients diagnosed with SNHI of unknown genetic cause after first-tier NGS testing.

**Results:** We identified bi-allelic *STRC* variants in 11 patients (11% diagnostic yield), including homozygous deletions, compound heterozygous deletions and conversions, and compound heterozygous SNVs and CNVs. In total, we detected *STRC* variants in 27 patients, with 81.6% of these variants occurring in patients with mild to moderate SNHI.

**Conclusions:** This study represents the first large-scale clinical investigation utilizing LRS technology for the genetic diagnosis of SNHI. Our study highlights the diagnostic capabilities of LRS in detecting complex variants within the *STRC* and advancing our understanding of the genetic etiology of SNHI that remains unresolved by conventional NGS.

**Graphical Abstract:** 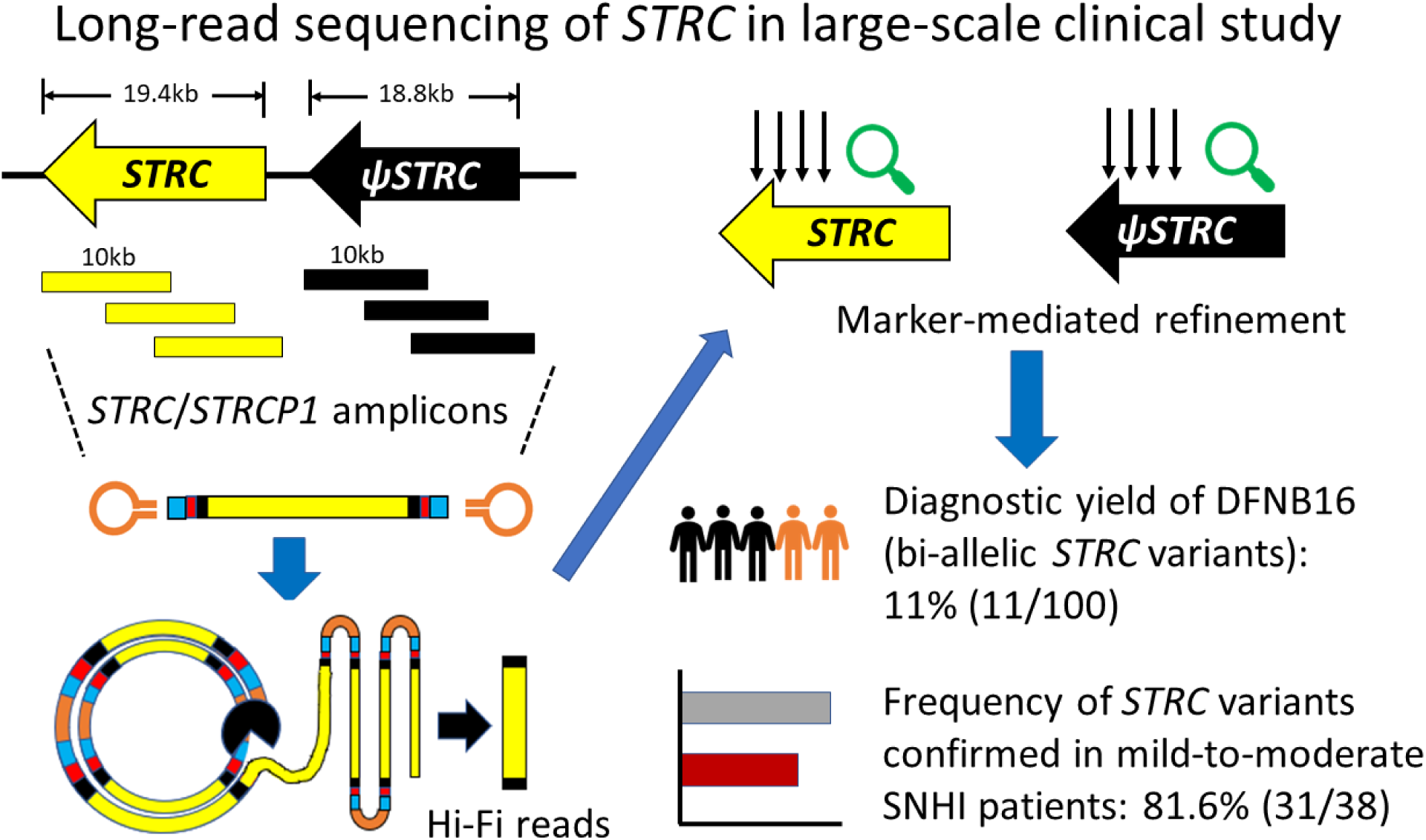

## Introduction

Sensorineural hearing impairment (SNHI) is a common childhood disorder with diverse etiologies. Genetic causes account for more than 50% of childhood SNHI cases and are classified as hereditary hearing impairment (HHI). To date, more than 120 genes have been associated with non-syndromic HHI [1, 2]. Among these, the stereocilin (*STRC*) gene, inherited in a recessive pattern (DFNB16, OMIM #603720), is known to cause non-syndromic HHI. The stereocilin protein encoded by *STRC* is expressed in the outer hair cell (OHC) bundles of the cochlea and is responsible for connecting adjacent stereocilia and interacting with the tectorial membrane [3]. This gene has been associated with mild-to-moderate SNHI and deafness-infertility syndrome, especially when long-range deletions occur across the *STRC* and *CATSPER2* region (15q15.3) [4, 5].

Pathogenic variants in *STRC* are a major contributor to HHI in different populations, with prevalence ranging from 5.6-10.2% in Europeans [6–8], 6.1-11.2% in Americans [9, 10], and 2.8-10.3% in East Asians [11–13]; studies have also confirmed *STRC* variants in Hispanic and Middle Eastern populations, with an overall prevalence of 16.1% in multi-ethnicity studies [14]. In addition to single nucleotide variants (SNVs), copy number variants (CNVs), also called structural variants (SVs), are important genetic causes of the *STRC*-related HHI. CNVs include a variety of large-scale genomic rearrangements ranging from 50 bp to megabases in size, such as deletions, duplications, translocations, and conversions [15]. CNVs have been identified in at least 29 HHI-related genes, with *STRC*, *OTOA*, and *GJB2*/*GJB6* being the most frequently implicated [16]. A meta-analysis by Han et al. [17], which included data from 37 relevant articles, showed that 14.4% of mild to moderate HHI patients have bi-allelic pathogenic *STRC* variants. Homozygous *STRC* deletions account for the largest proportion (∼70%) of these variants, highlighting their significant etiological role.

Another diagnostic challenge for *STRC* variants is the presence of the homologous pseudo-*STRC* (*STRCP1*) located ∼80kb upstream of the *STRC* region. *STRCP1* shares ultra-high sequence identity (97%) with *STRC* and has been implicated in gene-to-pseudogene conversion, complicating genetic testing [18]. While next generation sequencing (NGS) has demonstrated efficacy in identifying HHI-associated variants [2], its short-read nature (150-300 bp) limits its ability to comprehensively address both CNVs and pseudogenes simultaneously. This limitation hinders the accurate detection of SNVs and CNVs in the *STRC* “dark region”, which extends from the five-prime untranslated region to exon 18 and shows 99.8% identity between *STRC* and *STRCP1* (with less than 30 bp nucleotide differences in an approximately 11 kb region). To overcome this challenge, several bioinformatic tools and fluorescence-based approaches have been developed to complement NGS for CNV detection. These include CNV calling algorithms [19, 20], array comparative genomic hybridization (aCGH) [21–23], allelic-specific droplet digital PCR [9, 10], or multiplex ligation-dependent probe amplification (MLPA) [24, 25]. In addition, long-range (LR) PCR/nested PCR [8, 9] has been used to reduce pseudogene contamination and improve the accuracy of conventional NGS approaches. However, these methods often require the integration of multiple approaches and complex hierarchical pipelines, requiring the preparation of numerous primers specific for each exon or intron for subsequent validations. A one-round genetic assay that can quickly, accurately, and efficiently address the challenging questions posed by the *STRC* gene would be of significant clinical and academic value.

Long-read sequencing (LRS) assays offer a promising solution to these challenges by enabling direct sequencing of large genomic regions, ranging from kilobases to megabases. Two commercial platforms, Pacific Biosciences (PacBio) [26] and Oxford Nanopore Technologies (ONT) [27], offer high-resolution LRS for the detection of SNVs, indels, and CNVs [28]. In particular, PacBio’s high-fidelity (HiFi) sequencing via circular consensus sequencing (CCS) has demonstrated high accuracy rates for both SNVs (>99%) and CNVs (>95%) [29], which has been successfully used to implement direct LRS assays on 193 medically relevant genes, including the challenging *STRC*. However, the application of LRS-mediated CNV detection in large-scale clinical studies is still limited.

In this study, we use amplicon-based LRS technology to perform two-step target enrichment of *STRC* genomic segments, followed by generation of 10kb HiFi reads using the PacBio CCS pipeline to detect SNVs and CNVs hidden in the *STRC* gene. This study provides a potential solution to the challenges of *STRC* variant detection in clinical genetics.

## Materials and Methods

### Subjects

This study included 100 patients with SNHI of unknown genetic cause after first-tier NGS [30], including 73 with mild-to-moderate SNHI and 27 with severe-to-profound SNHI. Three National Institute of Standards and Technology (NIST) reference samples-HG001 (NA12878), HG002 (NA24385), and HG005 (NA24631)-were used for benchmarking [31], whose lymphoblastoid cell line DNAs were obtained from the Coriell Institute for Medical Research (Camden, NJ, USA). Genomic DNAs of patients were extracted using the MagCore® genomic DNA extraction kit (Cat. No. MGB400-03). This study was approved by the National Taiwan University Hospital (approval number 202012083RIND), and informed consent was obtained from all participants or their legal guardians.

### Long-read amplification of STRC

Prior to the implementation of LRS assays using single-molecule real-time (SMRT) technology, the recommended PacBio workflow (PN 101-921-300) for the construction of amplicon-based PacBio SMRTbell^®^ libraries was followed. A two-step PCR procedure was performed for each sample prior to SMRTbell library construction. In the first step, four pairs of 42 bp primers were designed, each consisting of a 25 bp targeted primer (**Table 1**) and a 17 bp M13 sequence connected at the 5 prime end (5’-GTAAAACGACGGCCAGT for forward and 5’-CAGGAAACAGCTATGAC for reverse). These primers, supplied by MB MISSION BIOTECH Co., Ltd. (Taipei, Taiwan), were capped with the 5’ amino modifier group C6 (5AmMC6) and used with the TaKaRa LA Taq® kit (code no. RR002A) to generate the initial PCR amplicons. In the second step, multiple pairs of asymmetric M13-barcoded forward/reverse primers (as specified in the PacBio workflow) and the KAPA HiFi HotStart ReadyMix (KK2602) were used to generate M13-barcoded amplicons.

**Table 1.**
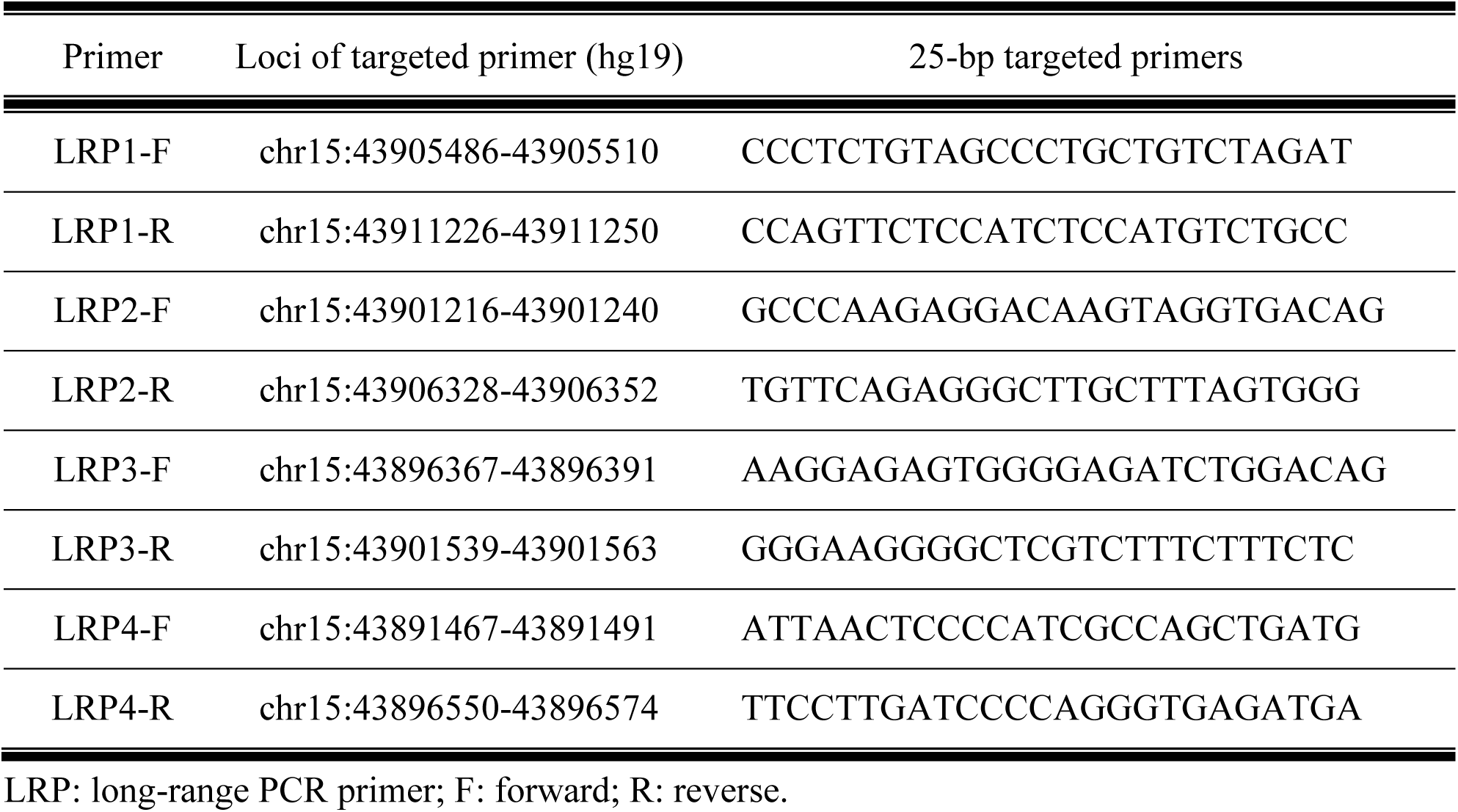
Targeted primers for amplifying *STRC* genomic regions.

These amplicons were then purified using AMPure® PB Beads and capillary electrophoresis, followed by SMRTbell library construction using the SMRTbell® Express Template Prep Kit 2.0 (PN 100-938-900). The SMRTbell products were then subjected to SMRT sequencing on the PacBio Sequel IIe platform using the Sequel II Sequencing Kit 2.0 (PN 101-820-200).

### Data analysis

Raw data from LRS assays were processed using SMRT^®^ Link (ver. 10.2.1, PacBio), the recommended software for analysis of PacBio sequencing data. Reads were mapped to the hg19 reference genome using pbmm2 (ver. 1.9) [32]. Variant calling, normalization, and annotation were performed using DeepVariant (ver. 1.4.0) [33], BCFtools (ver. 1.13) [34], and ANNOVAR (ver. 2021Oct19) [35], respectively. The merging and conversion of mapped BAM files was implemented using SAMtools (ver. 1.15.1) [36], and the refinement of long reads based on specific markers was conducted using Bamql (ver. 1.6) [37].

Multiple predictive scores were applied for the pathogenicity prediction of variants, including SIFT [38] and PolyPhen-2 [39] retrieved from dbNSFP (ver4.1) [40], CADD (GRCh37-ver. v1.7) [41], and SpliceAI (ver. 1.3.1) [42]. The allele frequencies of variants were retrieved from population databases gnomAD [43] (ver. 2.1.1, last accessed Oct 19, 2024) and Taiwan Biobank [44] (last accessed Oct 10, 2024). The pathogenicity assertions of variants were retrieved from disease databases ClinVar [45] (last accessed Oct 10, 2024) and Deafness Variant Database (DVD, ver. 9) [45].

### Multiplex ligation-dependent probe amplification (MLPA) assays

DNA samples (> 500ng) were prepared for validation of CNVs by MLPA assays. MLPA assays were performed using a commercial kit (SALSA^®^ MLPA^®^ Probemix P461-B1, MRC Holland, Amsterdam, The Netherlands). This experimental kit consisted of 45 MLPA probes within the chromosome 15q15.3 and 16q12.2 regions, including *STRC*, *CATSPER2*, *STRCP1*, *OTOA*, and other nearby regions. Seven and four probes in this kit were designed for the range of exons 19 to 28 on *STRC* and *STRCP1*, respectively. The experimental pipeline was based on the recommended general MLPA^®^ protocol (version-008).

## Results

### Target enrichment of the STRC region for generating 10kb amplicons

In this study, a total of 100 unrelated SNHI cases were recruited for the PacBio-mediated LRS assay, including 73 with mild-to-moderate SNHI and 27 with severe-to-profound SNHI. **Figure 1A** shows the amplicon-based target enrichment strategy for *STRC* using four pairs of PCR primers (see **Methods**) to generate 5.1-5.9kb amplicons. These amplicons, covering a quarter of the *STRC* region (19.4kb) and containing hundreds of base pairs of overlapping regions, were successfully amplified in the control samples. The amplicon length was further extended to 10kb using interlaced primer pairs, generating PCR products Amp-01, Amp-02 and Amp-03 in the control samples (**Figure 1B**). This approach also amplified homologous segments within the pseudogene *STRCP1*. The expected regions of the *STRC*/*STRCP1* amplifications are listed in **Table S1**.

**Figure 1.**
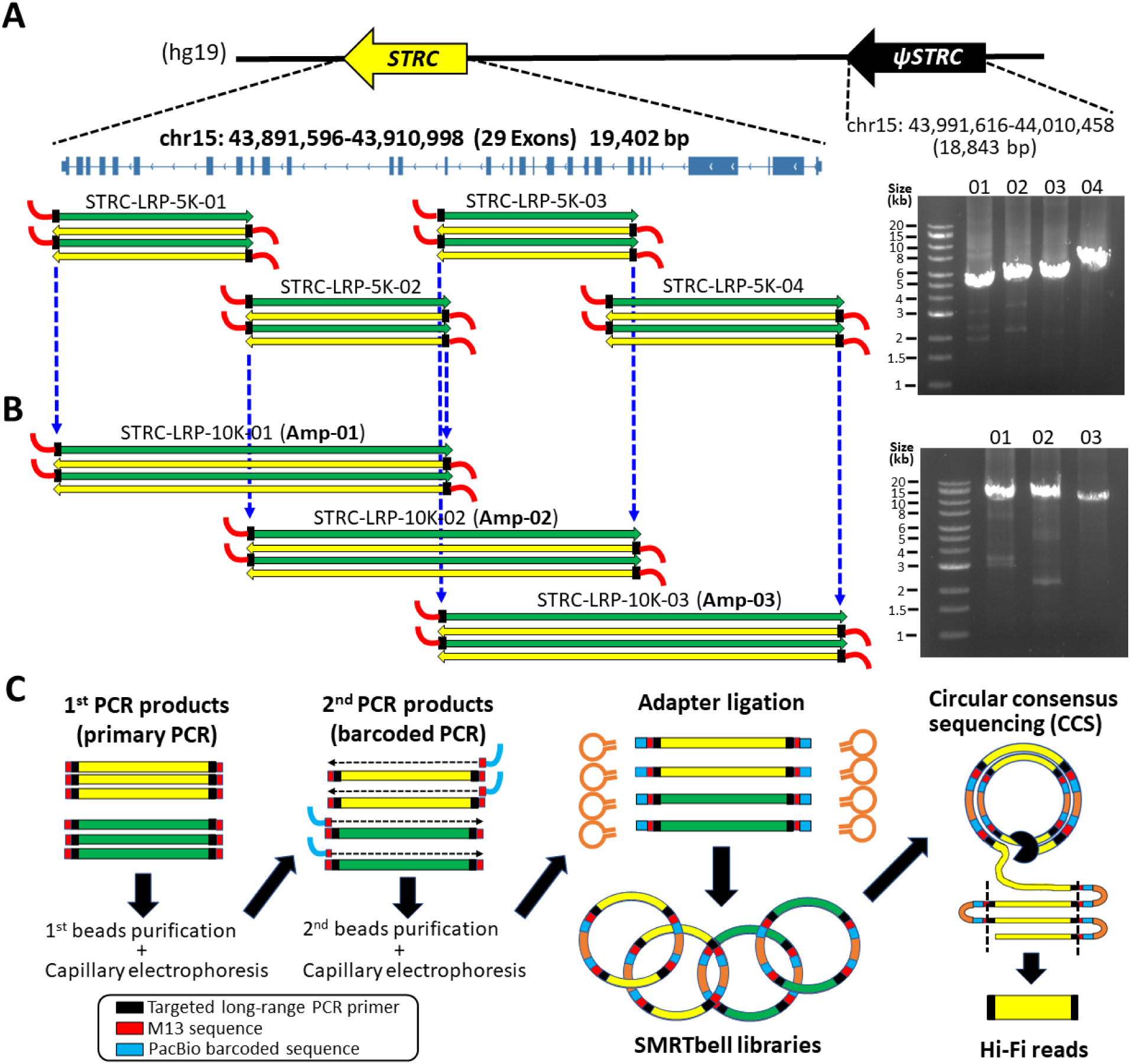
The workflow of *STRC* amplicon-based target enrichment, SMRTbell libraries, and circular consensus sequencing (CCS) for long-read sequencing (LRS). (A) The expected 5kb amplicons (STRC-LRP-5K-01 to -04) with their corresponding electrophoretic evidences. (B) The expected 10kb amplicons (STRC-LRP-10K-01 to -03, also referred to as Amp-01 to -03) with their corresponding electrophoretic evidences. (C) The two-step PCR of 10kb amplicons for SMRTbell library construction and CCS approach for Hi-Fidelity (Hi-Fi) reads.

SMRTbell libraries were constructed using these 10kb amplicons as circular templates for the PacBio CCS approach, generating HiFi sequencing data with 98-99% accuracy (**Figure 1C**). All samples, including three NIST reference samples (HG001, HG002, HG005), were mapped to the hg19 reference genome using PacBio-recommended pbmm2 software, followed by variant calling and annotation.

### Benchmarking and refinement of the STRC/STRCP1 copy number ratios using NIST reference samples

To establish benchmarks for our LRS assay, we first analyzed three NIST reference samples (HG001, HG002, and HG005). In addition to our LRS data,, we incorporated external sequencing data generated using the PacBio CCS pipeline (11kb) curated in the open access GIAB (Genome in a Bottle project) repository [46]. **Figure 2A** shows the mapping content of these reference samples on *STRC* and *STRCP1*. The distribution of mapped reads from the external source (upper panel) appeared symmetrical in HG001 and HG002, but asymmetrical in HG005. In contrast, our LRS assay showed an asymmetric distribution for HG001 and HG002 that was inconsistent with the external data (**Figure 2A**, bottom panel). This discrepancy suggested potential pseudogene contamination in our LRS assay.

**Figure 2.**
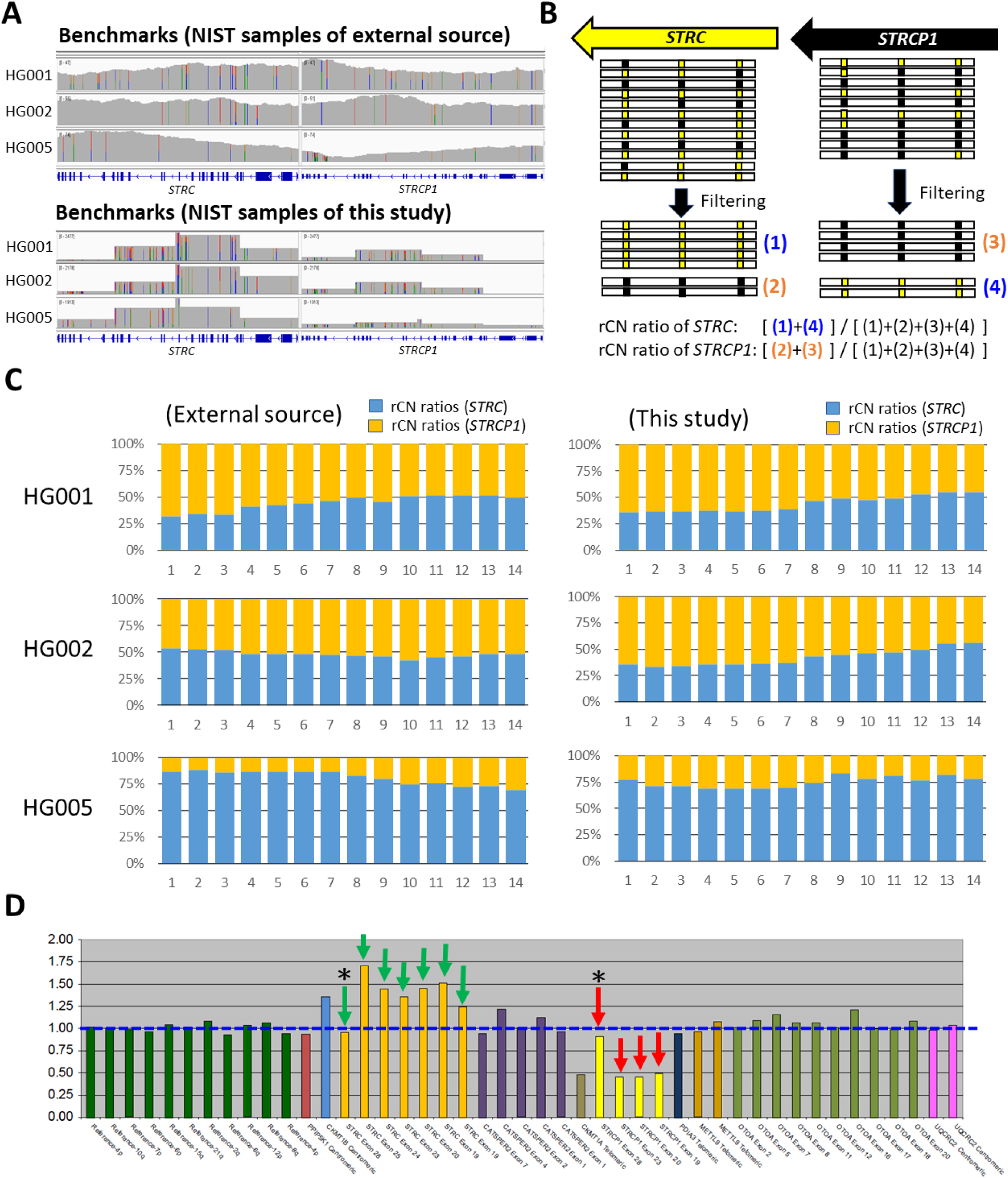
The refinement and validation of copy number ratios between *STRC* and *STRCP1* in benchmarks of NIST reference samples. (A) The overview of mapped LRS reads from external source and this study. (B) Illustrative processing of *STRC*/*STRCP1* refined copy number (rCN). (C) The rCN plots of reference samples (HG001, HG002, HG005) from external source and this study. (D) The MLPA plots of HG005. Green arrows: the MLPA signal peaks for *STRC*; red arrows: the MLPA signal peaks for *STRCP1*; y-axis: copy number compared to wild-type controls; blue dashed line: the baseline of normal copy number (copy number = 1.00). Asterisks indicate peaks directed at exon 28 of the *STRC*/*STRCP1* region. (Abbreviations) NIST: National Institute of Standards and Technology; MLPA: multiplex ligation-dependent probe amplification.

To address this, we developed a refinement process using a set of divergent markers between *STRC* and *STRCP1*, spanning from intron 26 to intron 15 (**Table 2** & **Figure S1**). These markers were grouped into 14 clusters in *STRC* (M1-M14) and *STRCP1* (pM1-pM14), respectively, based on their loci. To ensure accurate filtering of mis-mapped reads, these *STRC* markers, which represent the major differences between *STRC* and *STRCP1*, were selected based on their absence from the population genome (gnomAD database) with sufficient confidence (i.e., adequate coverage documented in the database). **Figure 2B** illustrates the filtering process, which produces four groups of filtered reads. Reads where all markers within a cluster at *STRC* and *STRCP1* contain the reference nucleotide (Ref) are categorized as group (1) and (3), respectively. Conversely, reads where all markers within a cluster at *STRC* and *STRCP1* match the alternative nucleotide (Alt) are classified as group (2) and (4), respectively. The refined copy number (rCN) ratios of *STRC* and *STRCP1* are then calculated as the sum of read counts in the combined group (1+4) divided by the sum of read counts in the combined group (1+2+3+4), and the sum of read counts in the combined group (2+3) divided by the sum of read counts in the combined group (1+2+3+4), respectively (**Figure 2B**, bottom panel).

**Table 2.**
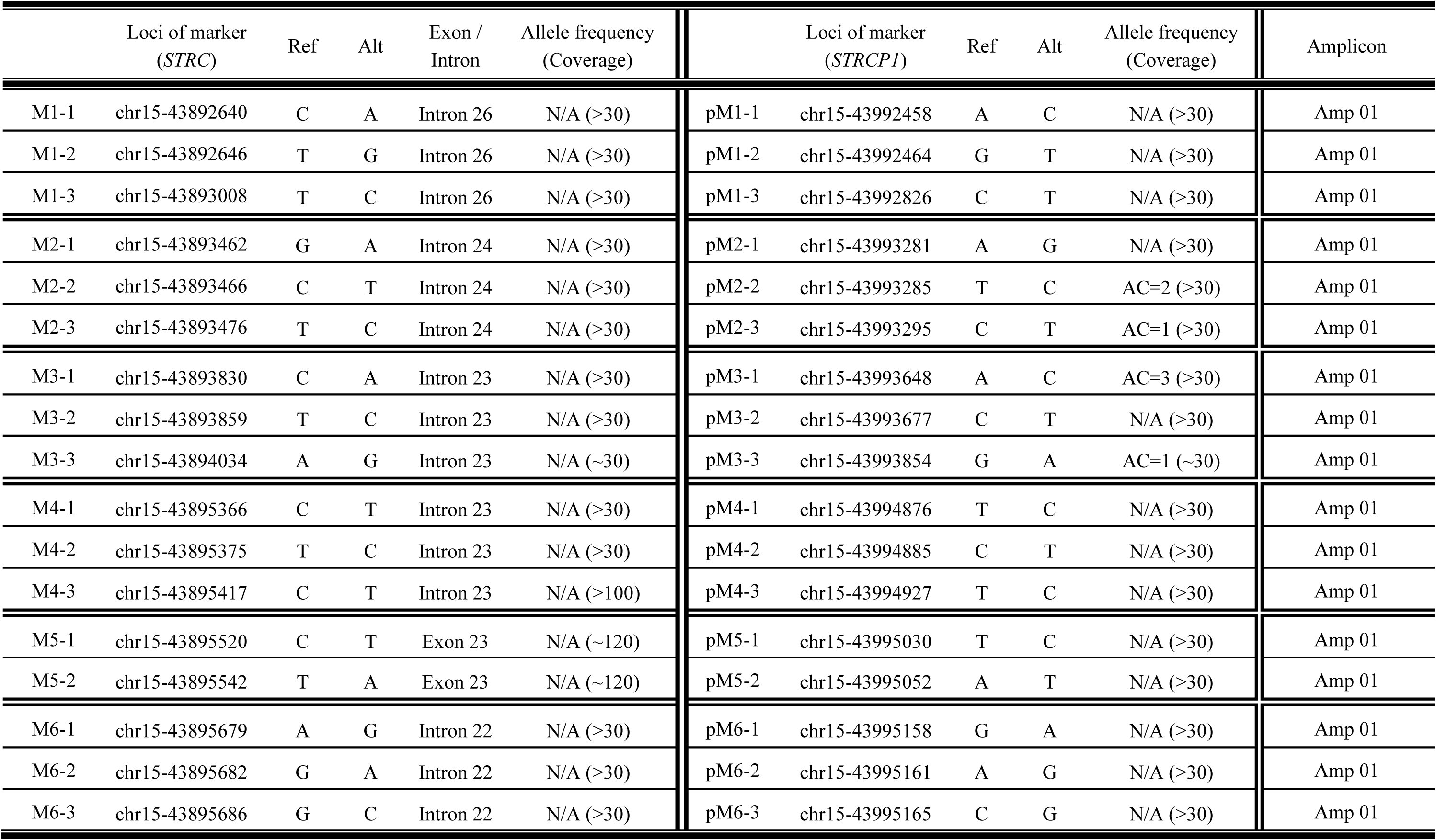
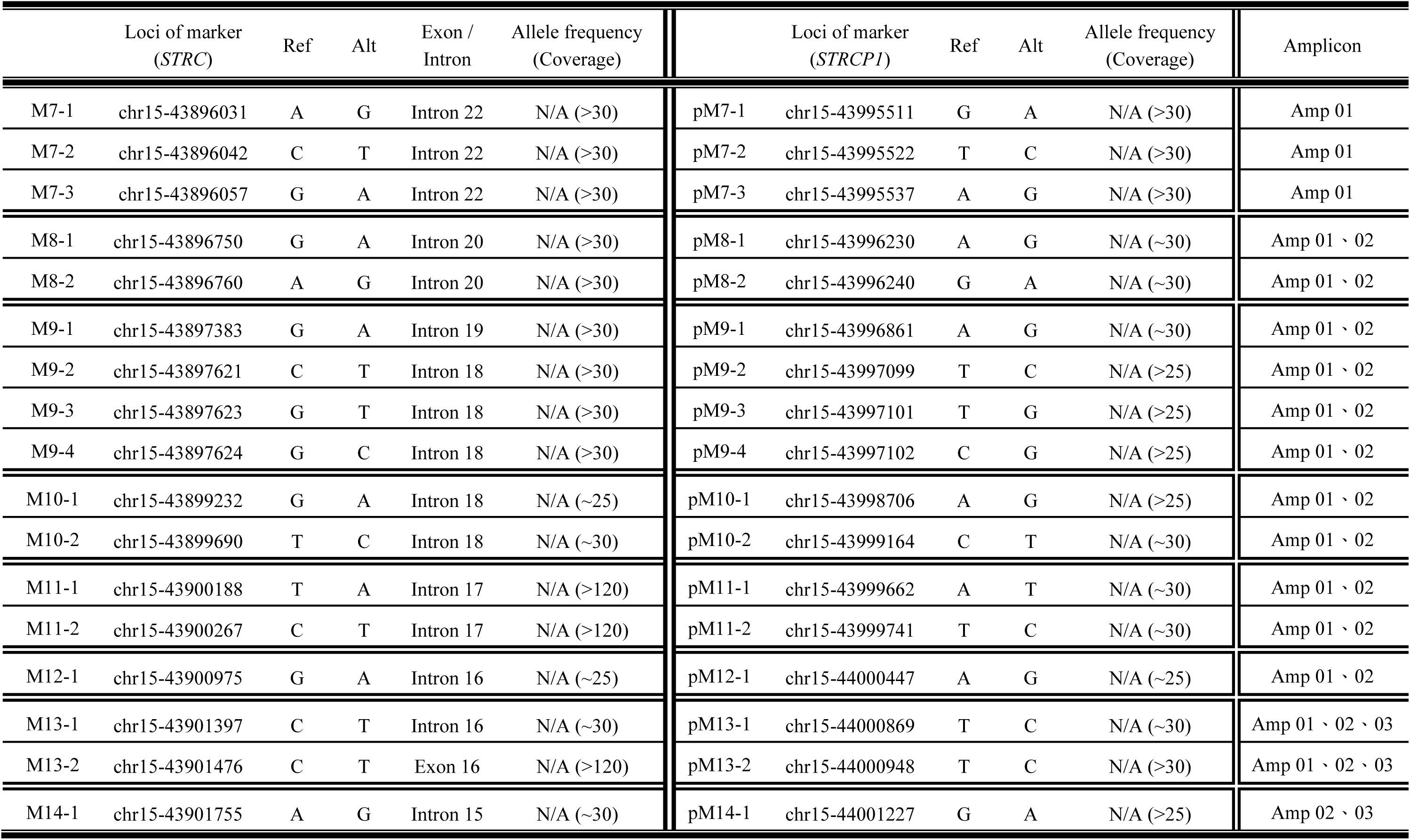
The 14 *STRC/STRCP1* divergent marker clusters.

Applying this rCN calculation (**Figure 2C**), HG001 and HG002 showed a similar trend of equal proportion per cluster between *STRC* and *STRCP1* in both the external source and our LRS data, supporting the feasibility of our refinement strategy. Interestingly, the *STRC* of HG005 showed a higher proportion in both datasets, suggesting a duplicated *STRC* in this sample. To confirm this finding, we performed MLPA analysis. As shown in **Figure 2D**, HG005 showed 1.5-fold MLPA signals at *STRC* and 0.5-fold signals at *STRCP1*, consistent with the rCN ratios and indicating a one-copy gain at *STRC* and one-copy deletion at *STRCP1* in HG005. Both HG001 and HG002 showed symmetrical *STRC*/*STRCP1* proportions in both rCN ratios (**Figure 2C**) and MLPA results (**Figure S2**), further validating the reliability of our refinement strategy.

### Marker-mediated refinement for SNV genotyping of STRC/STRCP1 in reference samples

In **Figure 2D**, the MLPA signal peaks on exon 28 of both *STRC* and *STRCP1* (arrows with asterisks) indicate an inconsistent copy number in contrast to the other signal peaks. This implies that there is an SNV present at this peak of *STRC* in HG005 that affects the trend of one-copy gain in the MLPA results. This SNV can be inferred as c.5125A>G (hg19:chr15:43892272-T-C, NM_153700.2:p.T1709A) because it is the only divergent nucleotide within exon 28 between *STRC* (chr15:43892272-T) and *STRCP1* (chr15:43992088-C) (**Figure S3**). This SNV is documented as “likely pathogenic” in ClinVar and is confirmed in the NIST external source data of HG005 but not of HG001 and HG002 (**Figure 3A**).

**Figure 3.**
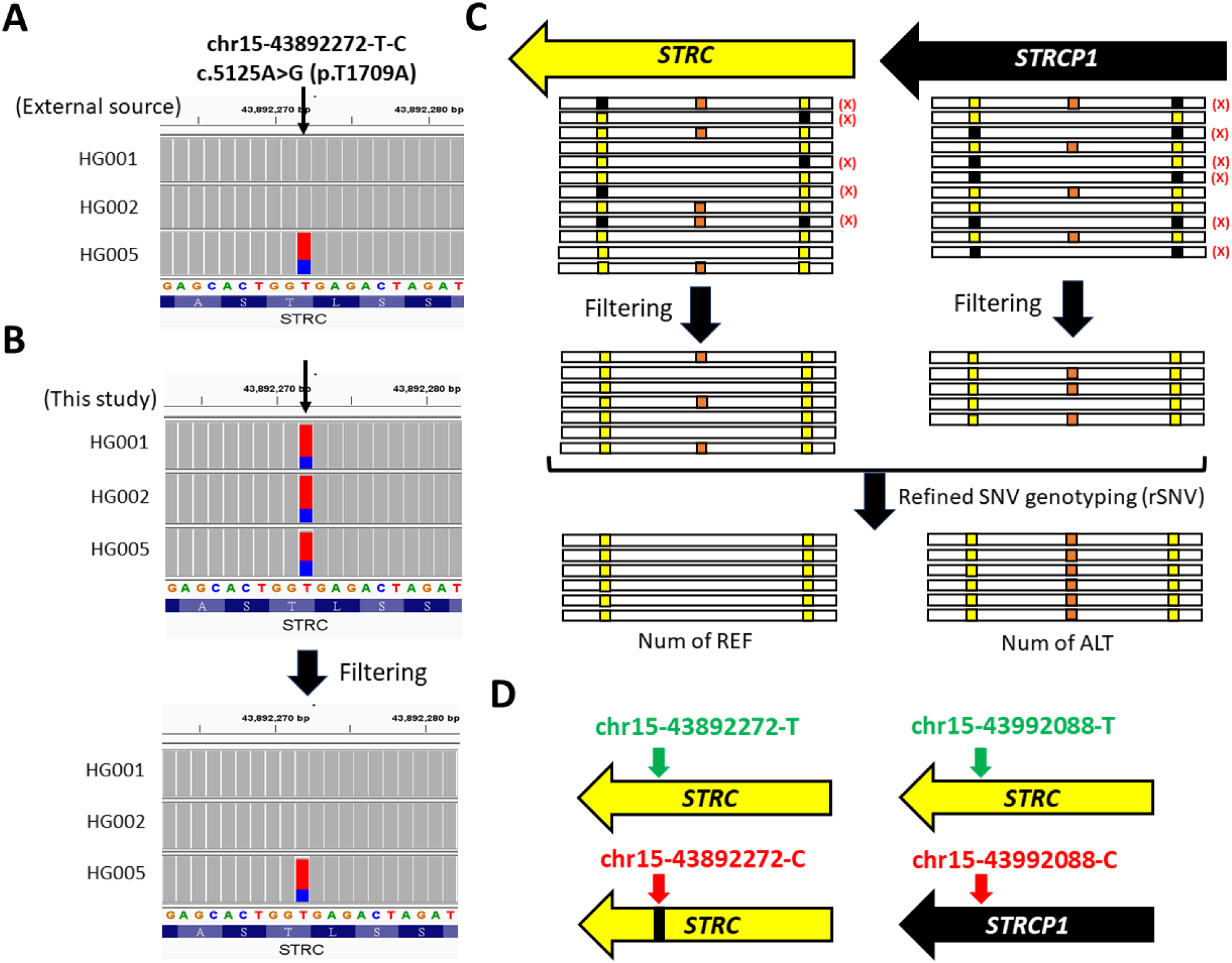
Marker-mediated refinement of single nucleotide variation (SNV) in reference samples. (A) Mapped reads from reference samples (NIST external source data). (B) Illustrative process of marker-mediated refinement for pre-filtered mapped reads resulting in refined SNV (rSNV) calls. (C) Illustrative process of marker-mediated refinement for accurate SNV genotyping of *STRC*/*STRCP1*. (D) Proposed *STRC*/*STRCP1* status of reference sample HG005.

However, our initial LRS assay mapping results indicated a heterozygous genotype of c.5125A>G in all three reference samples (top panel of **Figure 3B**), which is inconsistent with the NIST external source data. This discrepancy is likely due to pseudogene contamination. To address this, we applied a refinement process based on the *STRC/STRCP1* divergent markers (**Figure 3C**) to filter out mismapped segments using cluster M1, which is closest to the c.5125A>G variant. This refinement improved the mapping results for HG001 and HG002, while the result for HG005 remained unchanged (lower panel of **Figure 3B**). These refined results are consistent with the NIST external source data, demonstrating the effectiveness of marker-mediated refinement for accurate SNV genotyping.

Furthermore, the allele ratio of c.5125A>G in HG005 (NIST external source data) was 35% (Ref:Alt=40:22) rather than 50% (**Figure 3A**), reflecting the one-copy gain of *STRC*. Based on these results, we propose that reference sample HG005 harbors a one-copy gain of *STRC* with a heterozygous SNV c.5125A>G on *STRC* (**Figure 3D**). Therefore, we established benchmarks using NIST reference samples and applied marker-mediated refinement to our LRS results to ensure consistency with external source data.

### STRC variants (CNVs or SNVs) identified in HHI patients

Based on the marker-mediated refinement from reference sample benchmarks, both rCN ratios and refined SNV (rSNV) ratios of *STRC* are calculated for the detection of CNVs and SNVs, respectively, in each of the HHI patients included in this study. CNV analysis was performed using the following rCN ratio thresholds: two-copy loss (serial rCN ratios < 5% or mapping reads < 10 within *STRC* region), one-copy loss (serial rCN ratios < 30%), one-copy gain (serial rCN ratios > 70%). Samples with potential CNVs were further validated using MLPA assays.

For SNV detection, rSNV ratios were obtained by marker-mediated refinement using the closest marker cluster to each annotated variant. SNV types were defined as follows: homozygote (rSNV ratio > 80%), heterozygote (rSNV ratio 40-60%), and heterozygote with one-copy gain (rSNV ratio 30-35%).

Our analysis revealed bi-allelic *STRC* variants in 11 cases (**Figure 4A**), including homozygous deletion (n=7), compound heterozygosity of deletion and conversion (n=2), and compound heterozygosity of pathogenic SNV and deletion/conversion (n=2). In addition, 16 cases had mono-allelic *STRC* variants, including heterozygous deletion (n=2), conversion (n=2), and SNV (n=12). Analysis of allele frequencies (**Figure 4B**) showed that *STRC* deletions (n=19, 9.5% frequency in the entire cohort) were predominantly found in the mild-to-moderate HHI group (13%). Similarly, *STRC* conversions (n=5, 2.5% frequency) were also predominantly observed in the mild-to-moderate HHI group (2.7%). In contrast, *STRC* SNVs were detected with similar allele counts in both mild-to-moderate (n=8) and severe-to-profound (n=6) groups.

**Figure 4.**
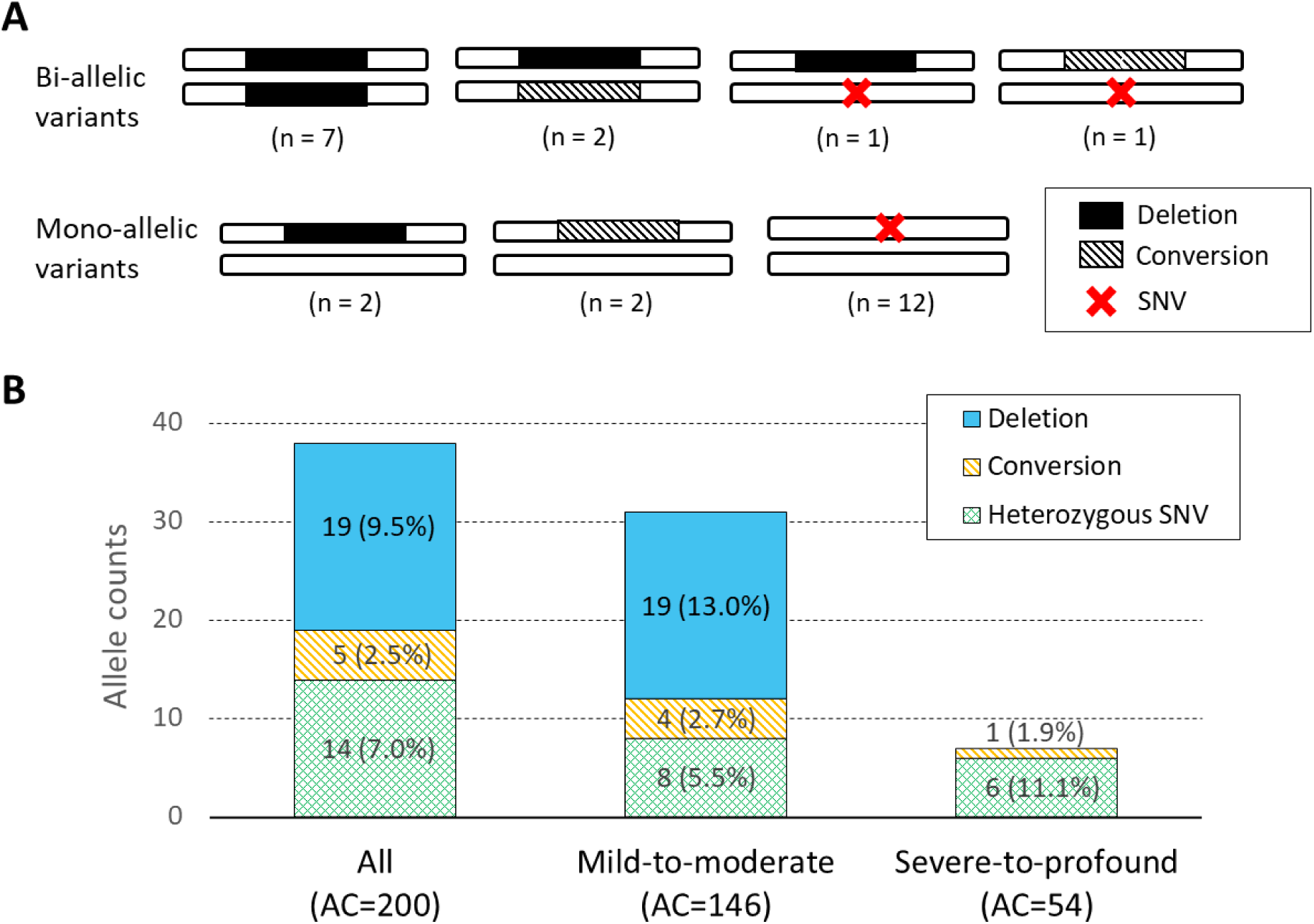
Statistical results of *STRC* variants found in the LRS assay. (A) Distribution of HHI cases with bi-allelic (n=11) and mono-allelic (n=16) variants. (B) Allele counts (AC) of identified CNVs and SNVs in all HHI cases and subgroups (mild-to-moderate, n=73; severe-to-profound, n=27).

Our LRS assays identified both 2-copy and 1-copy losses of *STRC*, as shown by the representative rCN ratio plots (**Figure 5**). These results, combined with MLPA validations, revealed several bi-allelic and mono-allelic CNVs, including deletions and gene conversions. Based on our LRS results, these *STRC* deletions and conversions spanned from 3’ end to intron 18 (markers M1 to M9) or to intron 15 (markers M1 to M14).

**Figure 5.**
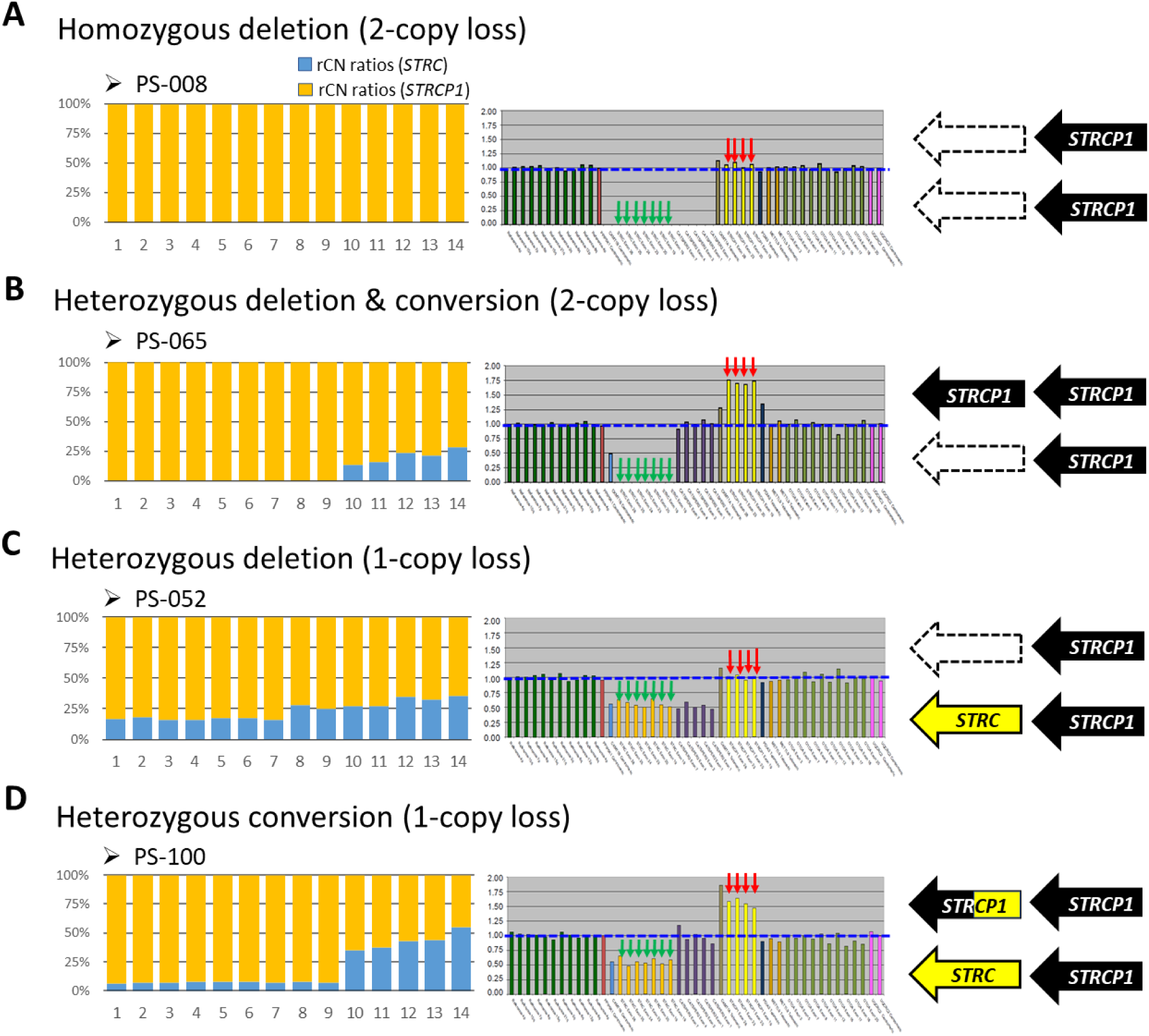
Four types of CNVs identified in this study, characterized by aberrant rCN ratios and MLPA peaks. (A) Homozygous deletions (PS-008 as an example); (B) Compound heterozygous deletion and conversion (PS-065 as an example); (C) Heterozygous deletion (PS-052 as an example); (C) Heterozygous conversion (PS-100 as an example).

In addition, we identified four pathogenic SNVs in *STRC* (**Table 3**). All SNVs were heterozygous with sufficient rSNV ratios for confident identification. These included c.5125A>G (n=9), c.4622G>A (n=3), c.4402C>T (n=1), and c.4143G>A (n=1). Of note, c.5125A>G (p.T1709A) represents the only divergent nucleotide between *STRC* and its pseudogene *STRCP1* reference transcript in exon 28 (i.e., the Alt nucleotide in *STRC* being as Ref nucleotide in *STRCP1*; see **Figure S3**). This variant is likely pathogenic based on predictive scores and ClinVar assertions (rs1336307815). The c.4622G>A (p.R1541Q) also shows likely pathogenicity according to predictive scores. In addition, we identified a previously reported pathogenic variant, c.4402C>T (p.R1468X), and a novel pathogenic variant, c.4143G>A (p.W1381X), both predicted to result in protein truncation.

**Table 3.**
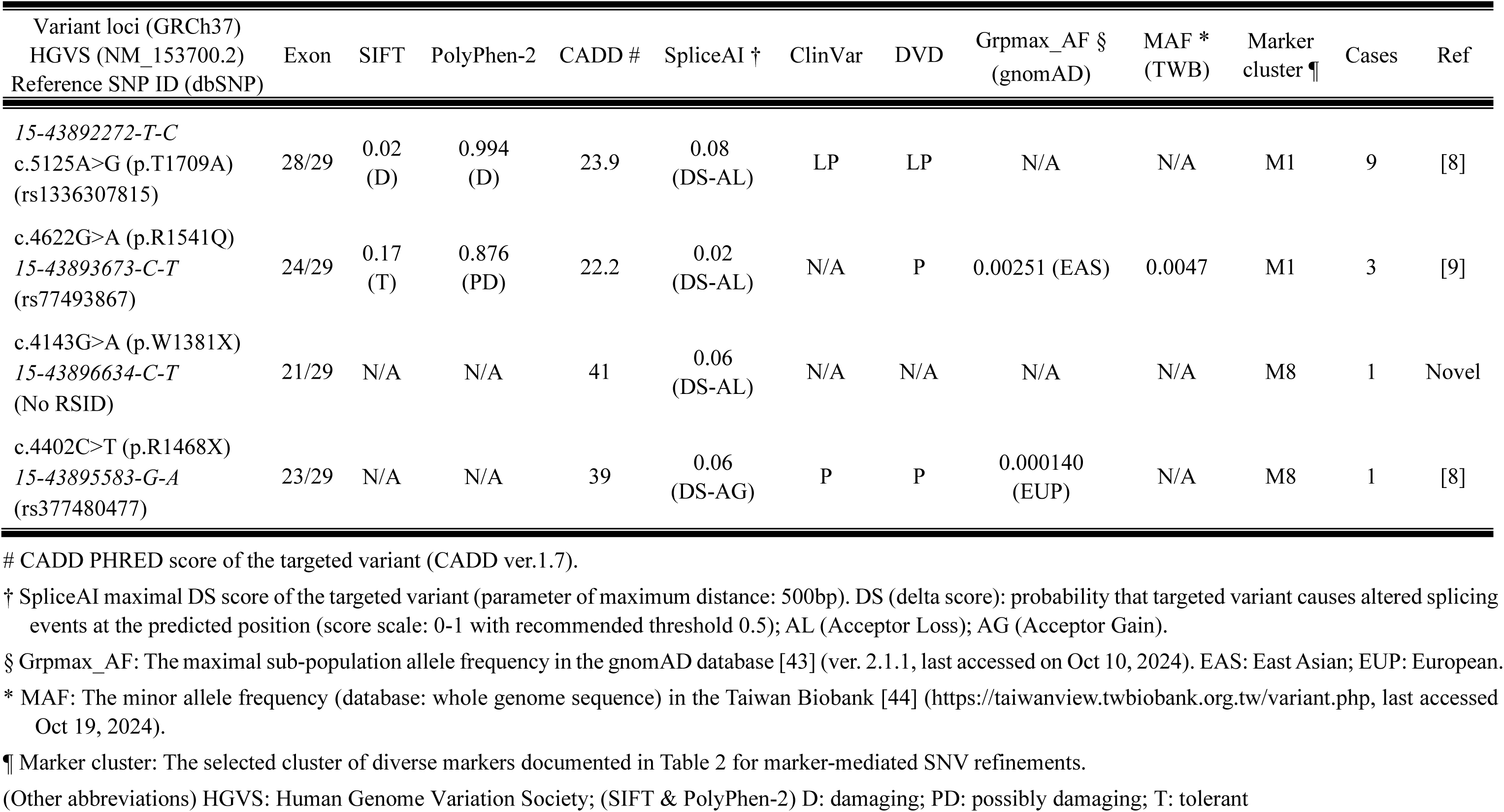
Pathogenic SNVs of *STRC* confirmed in this study.

### Compound heterozygosity of SNVs and CNVs in HHI cases

Among these HHI patients with confirmed *STRC* variants, we identified two individuals, PS-071 and PS-032, harboring compound heterozygous variants consisting of a SNV and a CNV. In patient PS-071, we observed homozygosity of the protein truncating variant c.4402C>T (**Figure 6A**). In addition, LRS and MLPA analysis revealed gene conversion spanning marker clusters from M1 to M12 in the LRS rCN ratio plot (**Figure 6B**). These findings confirm compound heterozygosity (c.[4402C>T];[conversion]) in PS-071.

**Figure 6.**
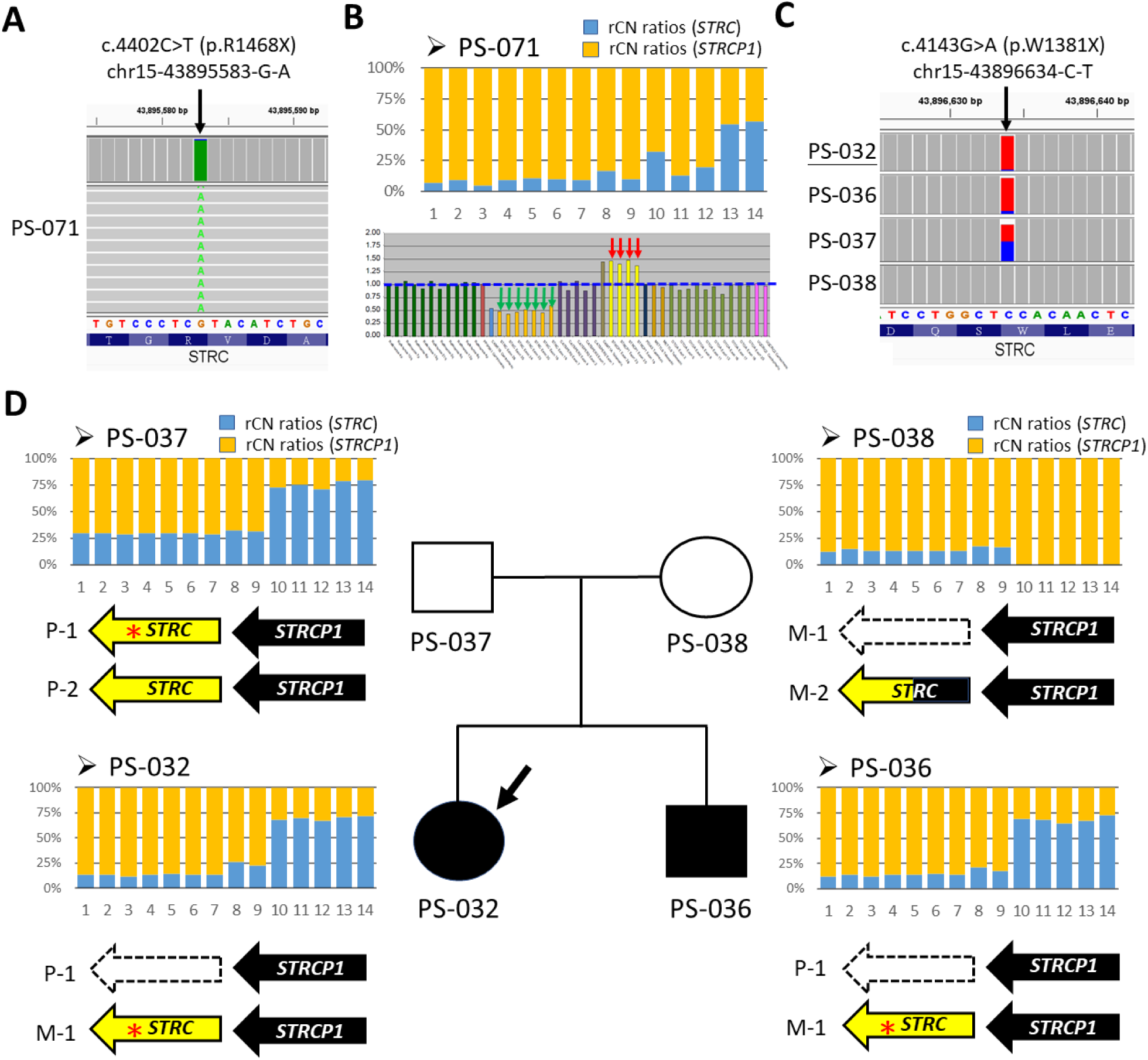
Compound heterozygous genotypes in two unrelated patients with truncating variants and either gene conversion or deletion. (A) c.4402C>T (p.R1468X) in PS-071 from a simplex family, forming a compound heterozygous genotype with (B) a single copy loss caused by gene conversion of *STRCP1*. (C) c.4143G>A (p.W1381X) in PS-032 and PS-036, inherited from the paternal allele, coupled with (D) a large deletion of *STRC*, inherited from the maternal allele, confirming compound heterozygosity in an autosomal recessive inheritance pattern.

Patient PS-032 was recruited from a multiplex family with an affected sibling (PS-036), an unaffected father (PS-037), and an unaffected mother (PS-038). A novel protein-truncating variant, c.4143G>A, was identified with homozygosity in both affected siblings and heterozygosity in the father (**Figure 6C**). Subsequent LRS and MLPA analysis revealed a heterozygous deletion spanning marker clusters from M1 to M9 in the LRS rCN ratio plot. This deletion, present in both affected siblings, was inherited from the maternal allele. (**Figure 6D** & **Figure S4**). These results confirmed the compound heterozygosity (c.[4143G>A];[deletion]) in both PS-032 and PS-036, indicating the autosomal recessive inheritance in this multiplex family.

However, in unaffected mother (PS-038), LRS analysis revealed missing rCN ratios for *STRC* from M10 to M14 (intron 15 to 18), a region beyond the detection range of the MLPA validation (**Figure S1**). Given the mother’s normal hearing phenotype, we conclude that she carries a benign partial gene conversion. Analysis of divergent nucleotides by *STRC*/*STRCP1* pairwise alignment (**Figure S5**) revealed only three exonic variants within the region spanning exons 1 to 18. All three are synonymous variants where the *STRC* sequence is directly replaced by the homologous *STRCP1* segment (**Table S2**), suggesting that this partial pseudogene conversion is likely harmless.

## Discussion

In this study, we used the amplicon-based LRS assays to investigate the genetic causes of *STRC*-related SNHI in 100 Taiwanese HHI patients unresolved by NGS-based diagnostics. Through marker-mediated refinement and MLPA validation, we identified disease-causing *STRC* variants, including CNVs and SNVs. Our LRS assay detected bi-allelic *STRC* variants in 11 patients, establishing a diagnostic yield of 11% for DFNB16 in this cohort. Considering both bi-allelic and mono-allelic variants, our results indicate a high allele frequency (19%) of *STRC* variants in 27 HHI patients. Notably, 81.6% of these variants were identified in patients with mild-to-moderate SNHI. This is the first study to report the genetic epidemiology of *STRC*-related SNHI using LRS, which demonstrated a prevalence of DFNB16 in the Taiwanese HHI cohort comparable to other East Asian populations [12, 47].

Previous studies across various populations have highlighted the significant contribution of pathogenic *STRC* variants to HHI [17]. In particular, NGS-based studies in Asian populations, complemented by CNV validation methods, have reported a high frequency of CNVs in patients with mild-to-moderate SNHI [12, 13]. While NGS effectively detects homozygous SNVs and deletions within the highly divergent region of *STRC* (exons 18-29) [6, 48, 49], the presence of the homologous pseudogene *STRCP1* and potential gene-to-pseudogene conversion events can complicate variant interpretation [18]. These challenges can lead to reduced coverage and hinder accurate variant annotation and analysis [50, 51]. Although MLPA is a widely used tool for CNV detection [6, 13, 52], it may not effectively capture the CNVs within the *STRC*/*STRCP1* highly-homologous region spanning from exons 1 to 18 [13, 51].

LRS technologies, with their ability to sequence long genomic regions (kilobases to megabases) [53], offer a potential solution for the comprehensive detection of both SNVs and CNVs [28, 54]. While earlier LRS methods suffered from high error rates (∼5-20% base calling errors)[55], recent advances in the PacBio and ONT platforms have significantly improved accuracy, with error rates now down to <1% [29] and <5% [56], respectively. This increased accuracy makes LRS suitable for clinical applications.

In this study, we used a PacBio-based LRS approach, following the recommended amplicon-based workflow, to decipher hidden *STRC* variants in a Taiwanese HHI cohort. Although pseudogene contamination in our LRS assays initially hindered variant interpretation, we developed a marker-mediated refinement strategy based on *STRC*/*STRCP1* divergent marker clusters to address it. This approach exploits the concept of genetic linkage [57], which suggests that proximal genetic loci are more likely to occur on the same haplotype. The performance of our LRS-based refinements has been validated using NIST reference samples, with results consistent with external sources and our own experiments.

For CNV detection, our refined copy number (rCN) ratios derived from the divergent marker cluster (M1 to M14) effectively addressed pseudogene contaminations and provided normalized results integrated with *STRCP1* screening data. Our rCN ratios also cover a broader range (intron 15 to exon 29) than conventional MLPA assays (exon 19 to exon 28). For SNV detection, the confirmed rSNVs in our study (**Table 3**) represent high-confidence genotyping in *STRC*. We also identified false-positive pathogenic variants resulting from pseudogene contaminations in our LRS assays due to low-confidence allele ratios of alternative nucleotides (**Table S3**). These variants are characterized by opposite REF/ALT nucleotides between *STRC* and *STRCP1*. Compared to the short reads of NGS, LRS offers the advantage of long-range sequence information, allowing the inclusion of more divergent markers near the targeted SNV for refinement.

To our knowledge, this is the first study to apply LRS technology to detect genetic causes in a large HHI cohort. Our study demonstrates the power of LRS in addressing challenging variants in *STRC* and provides valuable insights into the genetic etiology of HHI that remain unresolved by conventional NGS diagnostics. However, several limitations of this study deserve discussion. First, the limitations of long-range PCR, coupled with the recommended length of the CCS approach for HiFi reads, restricted amplicon generation to 10 kb instead of 20 kb, limiting the full coverage of the entire *STRC* region. Second, the lack of divergent markers in the highly homologous region spanning exons 1 to 15, which was previously reported to have nearly 100% identity between *STRC* and *STRCP1* [9], may hinder the refinement process for *STRC* variants within in this region. To improve clinical performance in the future, the long-range PCR workflow targeting the *STRC*/*STRCP1* divergence sites [8, 9] could be combined with amplicon-based LRS assays to reduce pseudogene contaminations.

## Conclusion

Our study highlights the diagnostic potential of LRS for detecting challenging variants in *STRC* and resolving the genetic etiology of HHI that remain unresolved by conventional NGS. The high allele frequency (19%) of *STRC* variants observed in this cohort emphasizes the importance of comprehensive *STRC* screening in HHI patients, especially those with mild-to-moderate SNHI. Future studies incorporating LRS-based *STRC* screening in clinical genetic testing are needed to further evaluate its diagnostic utility.

## Acknowledgements

We sincerely thank the A1 Laboratory of Genetic Testing of National Taiwan University Hospital (NTUH), GenePhile Bioscience Laboratory of Ko’s Obstetrics and Gynecology Clinic, Blossom Biotechnologies, Inc. (Pacific Biosciences distributor in Taiwan), and Taiwan Genome Industry Alliance Inc. (Taipei, Taiwan) for their invaluable experimental resources and technical support. We also thank all the subjects and their families for their generous contributions to this study.

## Ethics approval and consent to participate

All the patients and/or their families signed an informed consent form before participating in the study. All procedures used in the study were approved by the Research Ethics Committee of the National Taiwan University Hospital (201104025RC).

## Consent for publication

All patients have provided written informed consent.

## Data availability

The resultant datasets in this study are included within the article. Full datasets are available from the corresponding authors upon reasonable request.

## Authors’ contributions

Conceptualization: *Cheng-Yu Tsai, Pei-Lung Chen, Jacob Shu-Jui Hsu,* and *Chen-Chi Wu*

Investigation: *Cheng-Yu Tsai* and *Yu-Ting Chiang*

Validation: *Cheng-Yu Tsai* and *Yue-Sheng Lu* Formal analysis: *Cheng-Yu Tsai*

Visualization: *Cheng-Yu Tsai*

Resources: *Pei-Hsuan Lin, Chuan-Jen Hsu,* and *Pei-Lung Chen*

Project administration: *Cheng-Yu Tsai, Yue-Sheng Lu,* and *Ming-Yu Lo*

Funding Acquisition: *Shih-Feng Tsai, Jacob Shu-Jui Hsu,* and *Chen-Chi Wu*

Supervision: *Jacob Shu-Jui Hsu,* and *Chen-Chi Wu*

Writing (Original Draft): *Cheng-Yu Tsai,* and *Chen-Chi Wu*

Writing (Review/Editing): *Cheng-Yu Tsai, Pei-Lung Chen, Jacob Shu-Jui Hsu,* and *Chen-Chi Wu*

