## Supplementary Materials for "Comprehensive genetic analysis of *STRC* variants in hereditary hearing impairment using long-read sequencing"

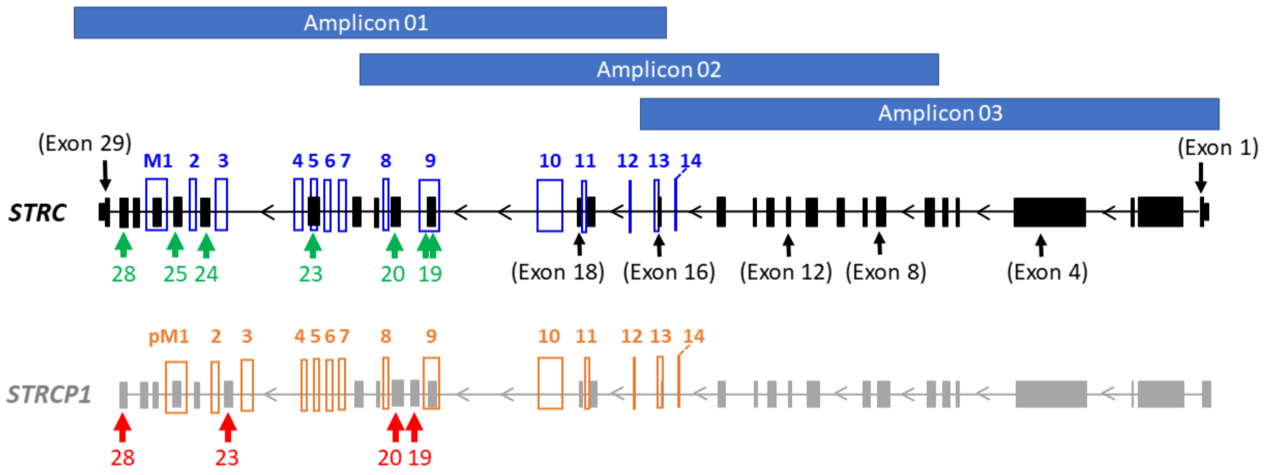


**Figure S1.** The overall locations of *STRC*/*STRCP1* divergent marker clusters on genomic regions and 10kb amplicons (Amplicon 01-03). The 14 marker clusters are used for calculating the refined copy number (rCN) ratios of *STRC* (blue boxes) and *STRCP1* (orange boxes). The probe locations of multiplex ligation-dependent MLPA assays in *STRC* (green arrows) and *STRCP1* (red arrows).

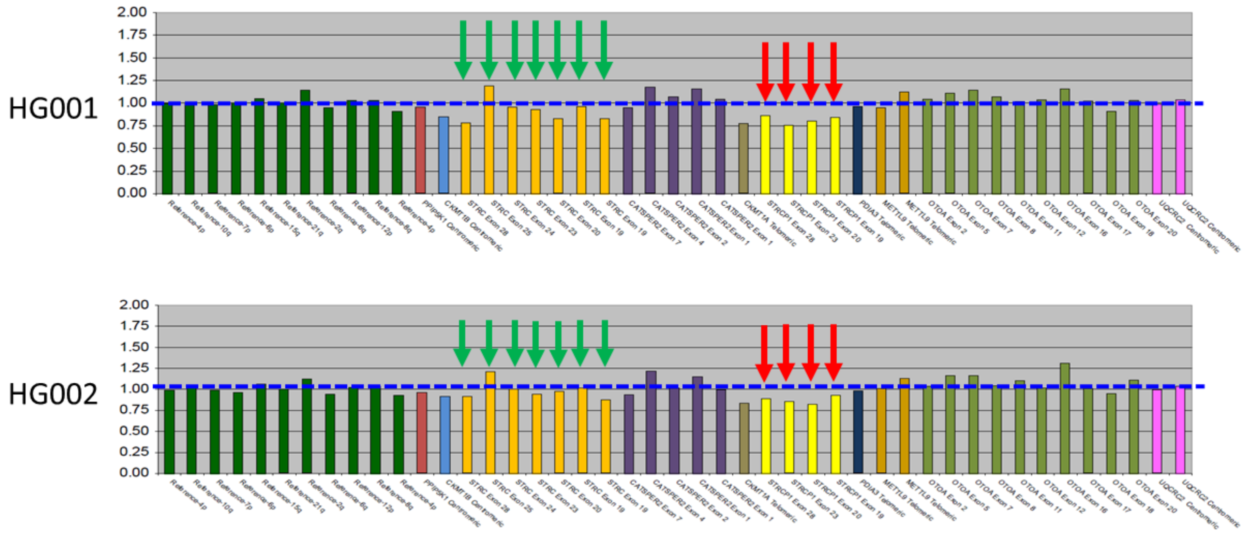


**Figure S2.** The MLPA plots on NIST reference samples HG001 and HG002. Seven probes (in orange) for *STRC* and four probes (in yellow) for *STRCP1* show the trend of normal copy number. Green arrows: the MLPA signal peaks for *STRC*; red arrows: the MLPA signal peaks for *STRCP1*; y-axis: copy number compared to wild-type controls; blue dashed line: the baseline of normal copy number (copy number = 1.00). (Abbreviations) NIST: National Institute of Standards and Technology; MLPA: multiplex ligation-dependent probe amplification.

```
43892158  CTTGCTGTTCTGGGCTCTCCTTTCCCTCATGTTGGGCCCATGCAACTGCT  43892207
>>>>>>>   ||||||||||||||||||||||||||||||||||||||||||||||||||  >>>>>>>
43991974  CTTGCTGTTCTGGGCTCTCCTTTCCCTCATGTTGGGCCCATGCAACTGCT  43992023

43892208  CGTCGCTGCTCAGGACTCAGAAAGGCCATTTGCTCAGGAGTGACAGCCAC  43892257
>>>>>>>   ||||||||||||||||||||||||||||||||||||||||||||||||||  >>>>>>>
43992024  CGTCGCTGCTCAGGACTCAGAAAGGCCATTTGCTCAGGAGTGACAGCCAC  43992073

43892258  AGCCTGAGCACTGGTGAGACTAGATAGTTGGATGGGACTAAACACCAC    43892305
>>>>>>>   ||||||||||||||.|||||||||||||||||||||||||||||||||    >>>>>>>
43992074  AGCCTGAGCACTGGCGAGACTAGATAGTTGGATGGGACTAAACACCAC    43992121
```

**Figure S3** Pairwise alignment of exon28 between *STRC* and *STRCP1*. The only divergent single nucleotide (in red box) is hg19:chr15-43892272-T (in *STRC*) and hg19:chr15-43992088-C (in *STRCP1*).

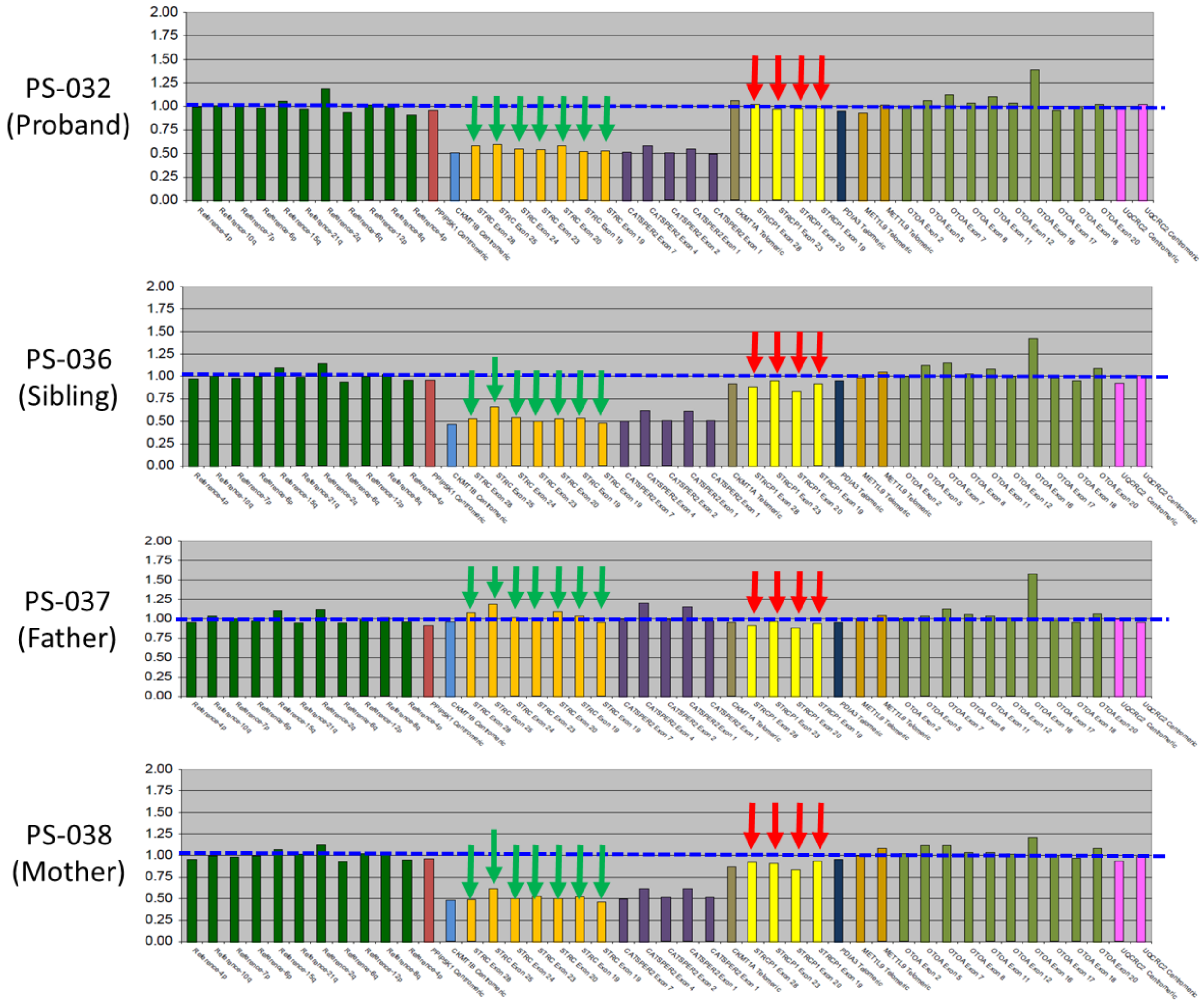


**Figure S4.** MLPA signal plots of the HHI multiplex family (PS-032, -036, -037, -038). A heterozygous deletion found in both proband (PS-032) and sibling (PS-036) was confirmed to be inherited from the maternal allele (PS-038).

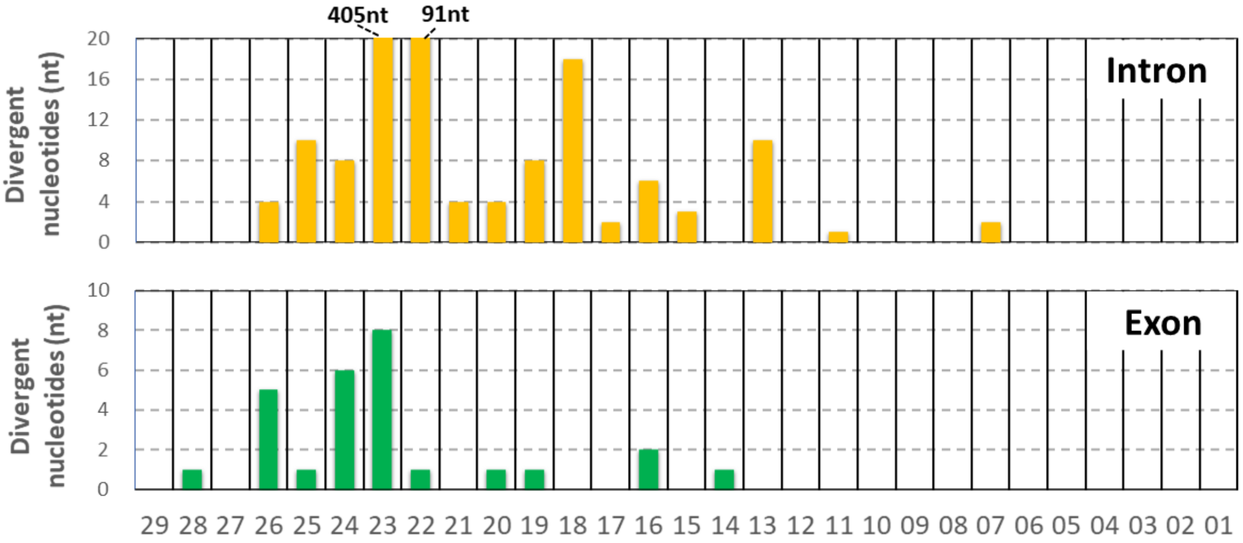


**Figure S5.** Statistical plot of *STRC*/*STRCP1* divergent nucleotides. The divergent content of each exon/intron is calculated by pairwise sequence alignment between *STRC* (NM_153700.2) and *STRCP1* (NR_146078.1).

**Table S1.** The expected regions of 10kb *STRC*/*STRCP1* amplicons

| Amplicon ID (PCR primers) | *STRC* (NM_153700.2) chr15: 43891761-43910998 | *STRCP1* (NR_146078.1) chr15:43991616-44010458 | Genomic regions (*STRC*) |
|---|---|---|---|
| Amplicon 01 (LRP4-F & 3-R) | chr15:43891467-43901563 (10,097 bp) | chr15:43991283-44001035 (9,753 bp) | 3'-UTR to Intron 15 |
| Amplicon 02 (LRP3-F & 2-R) | chr15:43896367-43906352 (9,986 bp) | chr15:43995847-44005814 (9,968 bp) | Intron 21 to Intron 06 |
| Amplicon 03 (LRP2-F & 1-R) | chr15:43901215-43911482 (10,268 bp) | chr15:44000687-44010944 (10,268 bp) | Intron 16 to 5'-UTR |

**Table S2.** Exonic variants within exon 1 to 18 caused by *STRCP1* conversion

| Variant locus (GRCh37) HGVS (NM_153700.2) | ClinVar | Grpmax_AF # | Number of Homozygotes # | Corresponding REF nucleotide of *STRCP1* |
|---|---|---|---|---|
| *chr15-43901476-C-T* Exon 16: c.3555G>A (p.L1185=) | LB | N/A | N/A | *chr15-44000946-T* |
| *chr15-43901491-A-C* Exon 16: c.3540T>G (p.L1180=) | B | 0.004051 (AFR) | 20 | *chr15- 44000983-C* |
| *chr15-43903129-A-G* Exon 14: c.3360T>C (p.C1120=) | B / LB | 0.02009 (ASJ) | 338 | *chr15-44002601-G* |

### "Grpmax_AF" and "Number of Homozygotes" were retrieved from database gnomAD (ver2.1.1).
(Abbreviations) AFR: African; ASJ: Ashkenazi Jewish

**Table S3.** False-positive *STRC* variants in this study

| Variant locus of *STRC* (GRCh37) HGVS (NM_153700.2) | Predictive scores # | Variant locus on *STRCP1* (GRCh37) | Applied marker cluster |
|---|---|---|---|
| *chr15-43892272-T-C* Exon 28: c.5125A>G (p.T1709A) | SIFT (D); PolyPhen-2 (D) CADD (23.9) | *chr15-43992088-C-T* | M1 |
| *chr15-43892822-C-A* Exon 26: c.4903G>T (p.V1635F) | SIFT (T); PolyPhen-2 (PD) CADD (22.2) | *chr15-43992640-A-C* | M1 |
| *chr15-43893673-C-T* Exon 24: c.4622G>A (p.R1541Q) | SIFT (T) PolyPhen-2 (PD) CADD (22.2) | *chr15-43993491-T-C* | M3 |
| *chr15-43896918-G-A* Exon 20: c.4057C>T (p.Q1353X) | CADD (42) | *chr15-43996398-A-G* | M8 |

### Both SIFT and PolyPhen-2 predictions were queried from Deafness Variation Database (ver. 9, https://deafnessvariationdatabase.org/); CADD scores were queried based on GRCh37-v1.7 database (https://cadd.gs.washington.edu/snv).

(Other abbreviations) HGVS: Human Genome Variation Society; (SIFT & PolyPhen-2) D: damaging; PD: possibly damaging; T: tolerant
